## Supplementary_materials for "Generalizing AI-driven Assessment of Immunohistochemistry across Immunostains and Cancer Types: A Universal Immunohistochemistry Analyzer"

**This file includes:**

Supplementary Methods

Supplementary Figs. 1 to 4

Supplementary Tables 1 to 5

References (1 to 4)

Supplementary Methods

Quantitative evaluation of F1 score at patch-level

At the patch level, the F1 score is used as an evaluation metric for cell detection. The F1 score is a popular metric for object detection in computer vision since it considers precision and sensitivity simultaneously. Its definition requires the number of true positives (TP), false positives (FP), and false negatives (FN).

$$F1 score=2 X\frac{precision X recall}{\left( precision+recall \right)}=\frac{\#TP}{\#TP+0.5 X (\#FP+\#FN)}$$

Since these metrics have been developed for the classification task, it is not obvious how to measure them in the context of object detection. Therefore, a hit criterion is defined as follows. For each cell class, we determine the TP, FP, and FN with the following process,

1. Sort cell predictions by their confidence score.

2. Starting from a cell prediction with the highest confidence score, check whether any ground-truth cell is within a valid distance (25 pixels in 0.19 microns per pixel (MPP)) from the cell prediction.

2-1. If there is no ground-truth cell within a valid distance, the cell prediction is counted as an FP.

2-2. If there are one or more ground-truth cells within a valid distance, the cell prediction is counted as TP. The nearest ground-truth cell is matched with the cell prediction and ignored from the further process.

3. Go back to 2. until the cell prediction with the lowest confidence score is reached.

4. The remaining ground-truth cells that are not matched with any cell prediction are counted as FN.

After the above process, we aggregate the total number of TP, FP, and FN per cell class over all samples; then, we compute the per-class F1 score. The mean F1 (mF1) score across cell classes (TC- and TC+) is used as the final score. Each result has been reproduced *×*30 times using Monte-Carlo dropout at test time, an algorithm developed to study the robustness of a deep learning model over small variations^1^. In Fig. 1, for each test set we compare the statistical significance (p-value *<* 0*.*05) between the best single-cohort model (left of the dotted line) with all other multiple-cohort models. *p*-value has been calculated using the Wilcoxon signed-rank test implemented by *scipy.stats*^2,3^.

Quantitative evaluation of tumor proportion score at slide level

Tumor proportion score (TPS) is calculated by the following equation:

$$TPS=100 X \frac{\#TC+}{\#TC- + \#TC+}$$

Programmed Death-Ligand 1 (PD-L1) expression is subgrouped according to the TPS cutoff 1% and 50%, i.e., classified into TPS < 1%, 1% ≤ TPS < 50%, and TPS ≥ 50%. For simplicity, TPS has been used as a general whole slide image (WSI)-level metric to compare all IHC quantification across AI models. For each WSI, three board-certified pathologists assign a TPS score following the official protocol^4^. A category (TPS < 1%, 1% ≤ TPS < 50%, or TPS ≥ 50%) is then assigned to the slide by applying the cutoff, for example for PD-L1 Lung. The WSI-level ground truth was based on the consensus of three board-certified pathologists. For example, if two pathologists determined a WSI as TPS<1% while one pathologist determined it as TPS 1-49%, the WSI was assigned to TPS<1%.

As explained above, we compute TPS using our model for each WSI, and compare it with the manually assigned one. Fig. 2 shows the standard accuracy computed as follows, given *N* number of WSIs in a test set (e.g., HER2 Breast), *y* and ˆ*y* are respectively ground truth (GT) and predicted category.

$$Accuracy=\frac{1}{N}\sum_{i=1}^{N} [yi = yˆi]$$

**Supplementary Fig. 1. Patch-level quantitative analysis of the artificial intelligence (AI) models sorted by cancer types.** H-Br, HER2 of breast; P-L, PD-L1 22C3 of lung; P-Br, PD-L1 22C3 of breast; P-LBlBr, PD-L1 22C3 of lung, bladder, and breast; PH-B, PD-L1 22C3 and HER2 of breast; PH-LBr, PD-L1 22C3 and HER2 of lung and breast; PH-LBlBr, PD-L1 22C3 and HER2 of lung, bladder, and breast.

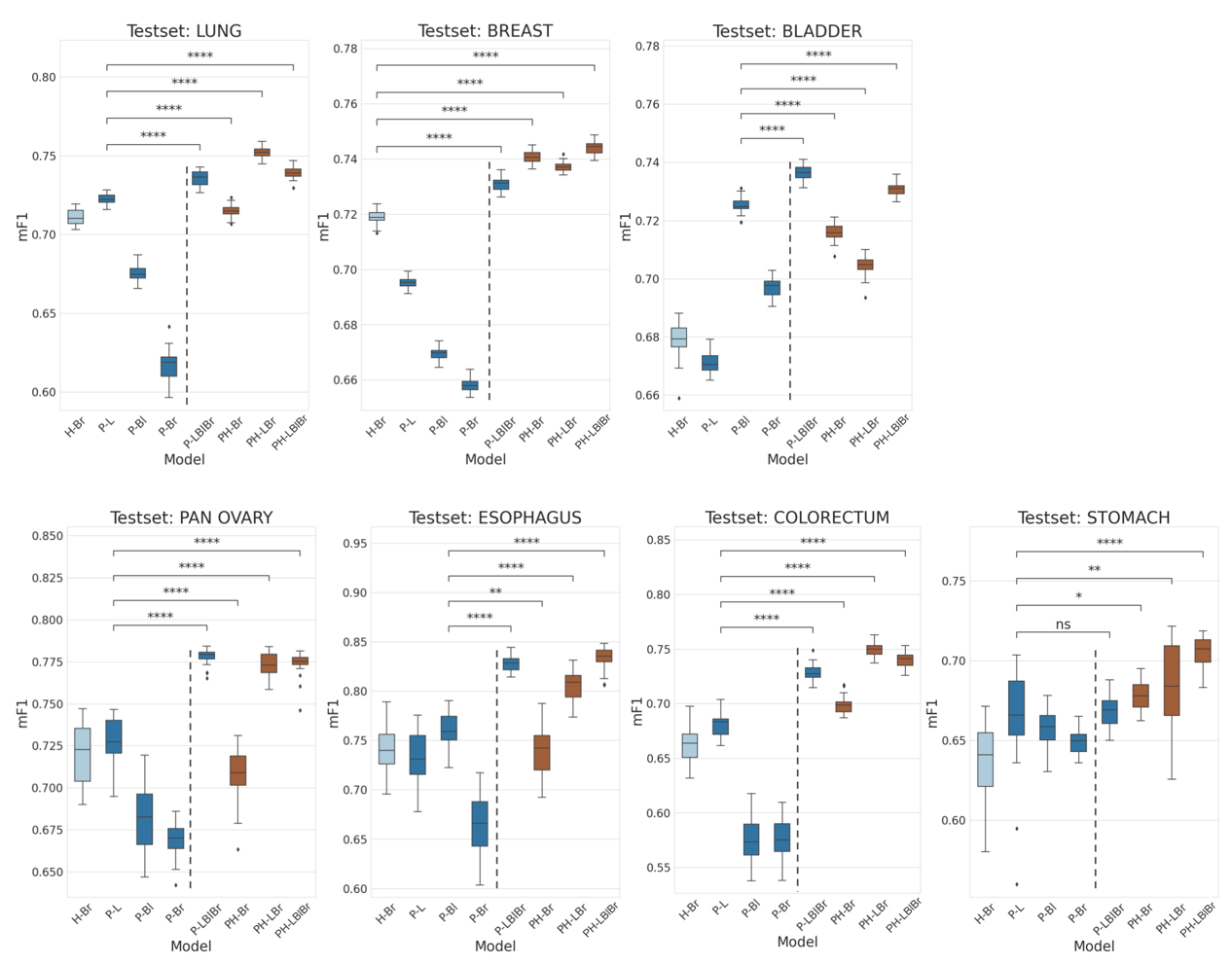

**Supplementary Fig. 2.** **Whole slide image (WSI)-level accuracy analysis of the artificial intelligence (AI) models.** **a** Macro-averaged accuracy of the eight AI models over all the stains. **b** Accuracy of the AI models in PD-L1 22C3 Lung dataset. **c** Accuracy of the AI models in PD-L1 22C3 Pan-cancer dataset. **d** Accuracy of the AI models in PD-L1 SP142 Lung dataset. **e** Accuracy of the AI models in multi-stain Pan-cancer dataset. The X-axis presents the summation of utilized stain types and the organ types of each cohort when training. Accuracy metrics are presented for 3-classes whole slide image (WSI) evaluation based on tumor proportion score (TPS) cutoffs. H-Br, HER2 of breast; P-L, PD-L1 22C3 of lung; P-Br, PD-L1 22C3 of breast; P-LBlBr, PD-L1 22C3 of lung, bladder, and breast; PH-B, PD-L1 22C3 and HER2 of breast; PH-LBr, PD-L1 22C3 and HER2 of lung and breast; PH-LBlBr, PD-L1 22C3 and HER2 of lung, bladder, and breast.

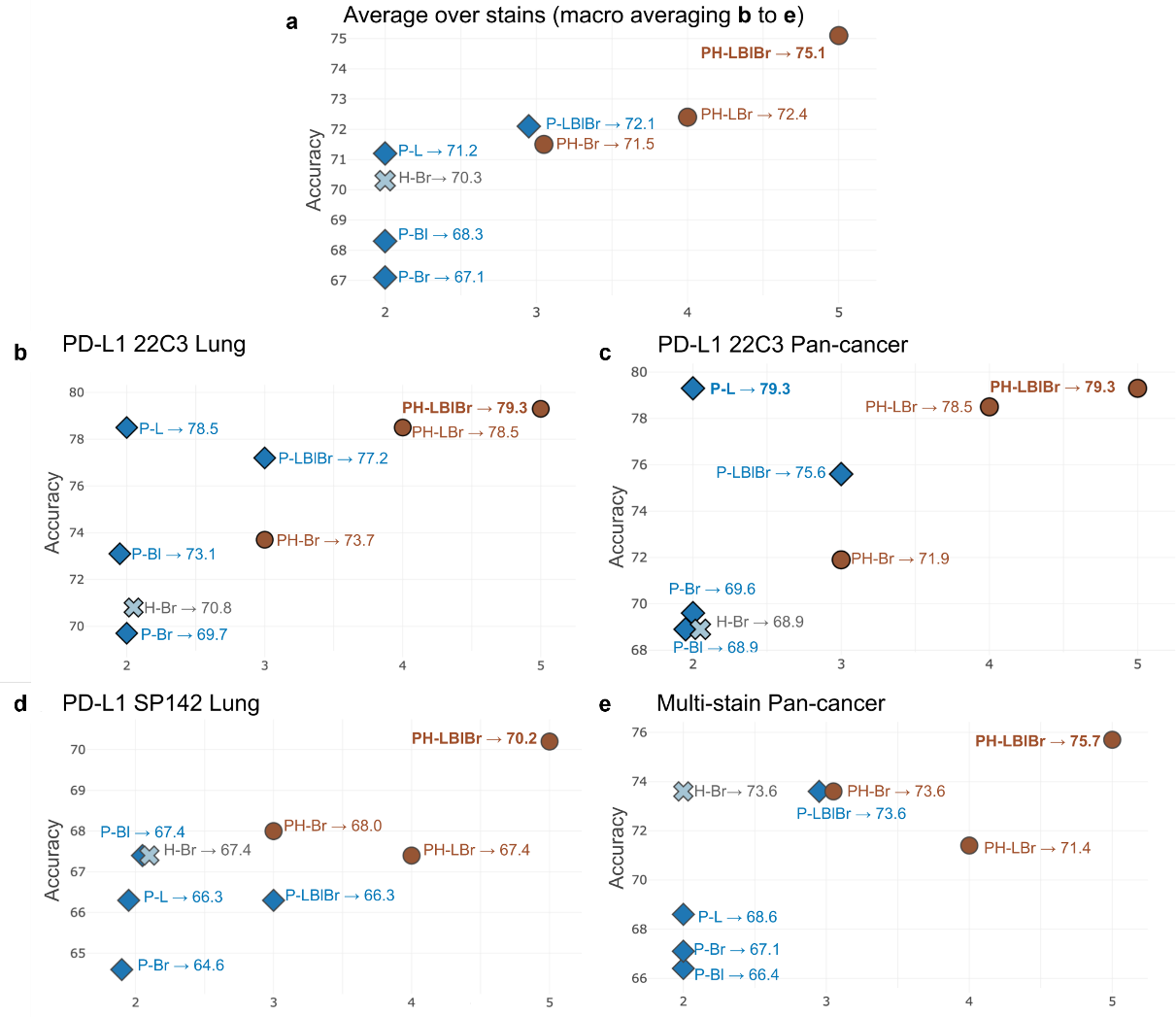

**Supplementary Fig. 3. Examples of whole slide images (WSIs) and their tumor proportion score (TPS) from training cohorts. a** Samples from training cohorts. **b** Samples from novel cohorts.

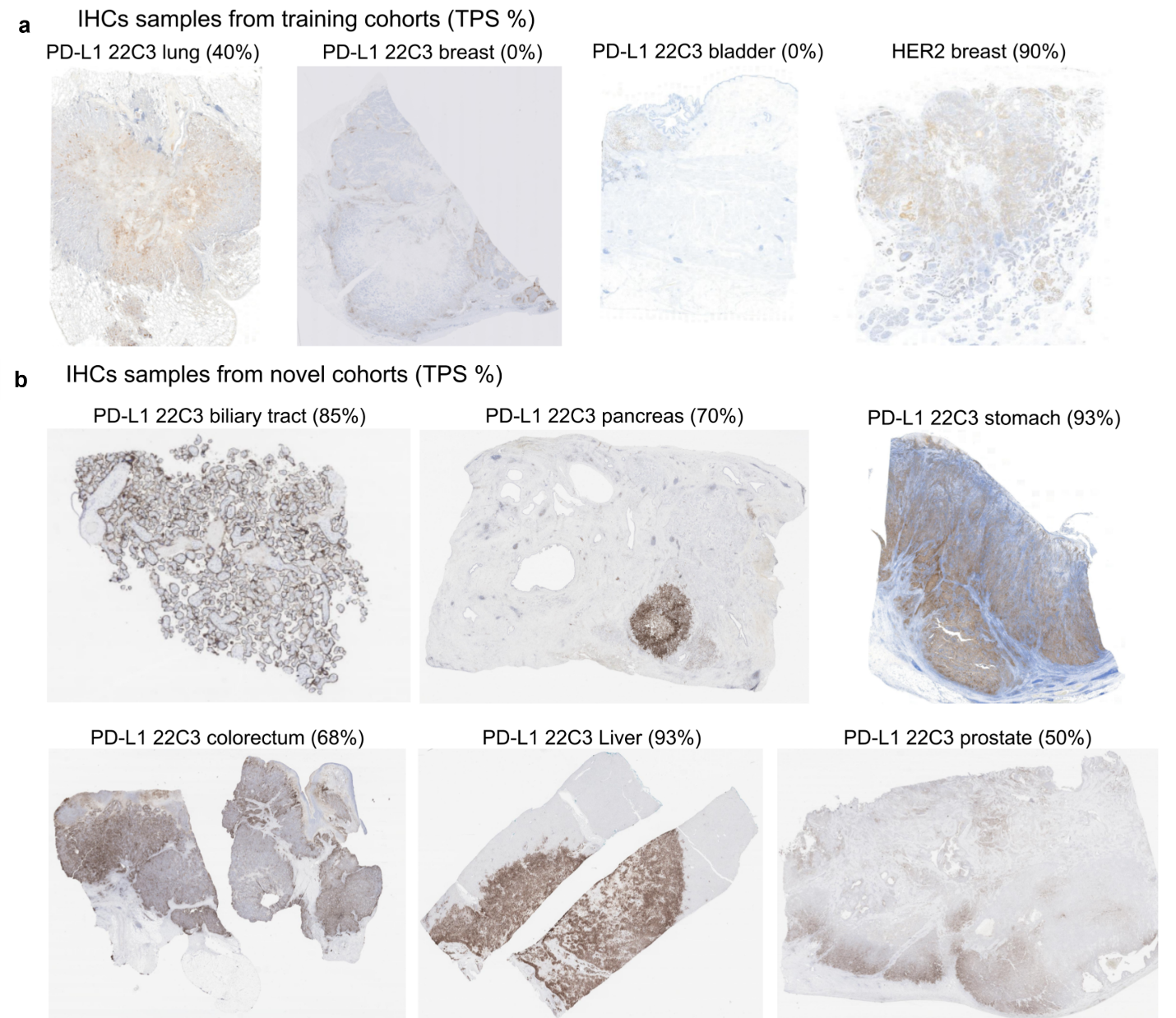

**Supplementary Fig. 4. Examples of whole slide images (WSIs) and their tumor proportion score (TPS) from novel cohorts.**

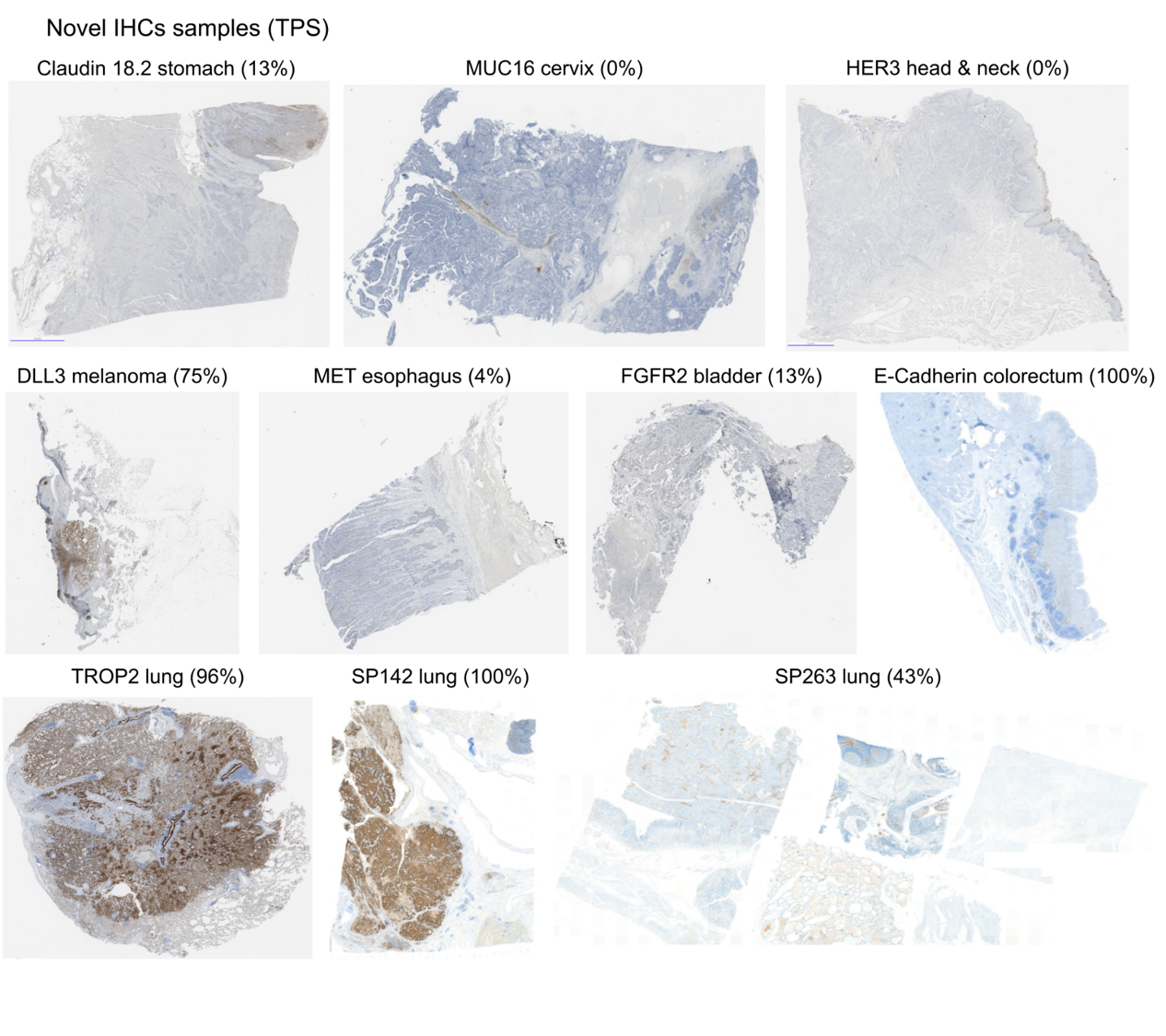

Supplementary Table 1. Number of slides for each whole slide image (WSI) test sets, divided by 1%/50% tumor proportion score (TPS) cutoffs into three classes.

| Dataset | TPS < 1% | TPS 1 − 49% | TPS ≥ 50% |
| --- | --- | --- | --- |
| PD-L1 22C3 Lung  (n = 479) | 81 (16.9%) | 162 (33.8%) | 236 (49.3%) |
| PD-L1 22C3 Pan-cancer  (n = 135) | 95 (70.4%) | 25 (18.5%) | 15 (11.1%) |
| PD-L1 SP142 Lung (n = 178) | 81 (45.5%) | 71 (39.9%) | 26 (14.6%) |
| Novel, multi-stain  (n = 140) | 71 (50.7%) | 37 (26.4%) | 32 (22.9%) |
| Total  (n = 932) | 328 (35.2%) | 295 (31.7%) | 309 (33.2%) |

Supplementary Table 2. Dataset configuration of the whole slide images (WSIs) used to develop the universal immunohistochemistry (UIHC) model.

| **Dataset** | **Source** | **Training** | **Tuning** | **Internal test** | **Total** |
| --- | --- | --- | --- | --- | --- |
| PD-L1 22C3 lung | Cureline | 329 | 41 | 0 | 370 |
|  | Aurora Dx | 381 | 32 | 50 | 463 |
|  | SNUBH | 0 | 28 | 29 | 57 |
|  | SMC | 0 | 62 | 49 | 111 |
| PD-L1 22C3 urothelial carcinoma | Cureline | 281 | 79 | 40 | 400 |
| PD-L1 22C3 Breast cancer | Cureline | 281 | 83 | 40 | 404 |
| HER2 4B5 Breast cancer | Cureline | 392 | 119 | 58 | 569 |
|  | AuroraDx | 415 | 118 | 60 | 593 |
|  | Superbiochips | 60 | 13 | 6 | 79 |
| Total |  | 2,139 | 575 | 332 | 3,046 |

PD-L1, Programmed Death-Ligand 1; HER2, Human Epidermal growth factor Receptor 2; SMC, Samsung Medical Center; SNUBH, Seoul National University Bundang Hospital

Supplementary Table 3. Antibody information for immunostains.

| Target | Antibody | Manufacturer | Dilution factor | Stain localization |
| --- | --- | --- | --- | --- |
| Claudin 18.2 | Anti-Claudin18.2 antibody [EPR19202] ab222512 | abcam | 1:50 | Membrane |
| DeLta-Like 3 (DLL3) | DLL3 (SP347), 08416931001 | Ventana | RTU | Membrane, Cytoplasm |
| E-cadherin | E-CAD-L-CE / Leica | Leica | 1:100 | Membrane |
| Fibroblast Growth Factor Receptor 2 (FGFR2) | Anti-FGFR2 antibody [SP273] – N-terminal ab227683 | abcam | 1:100 | Membrane, Cytoplasm |
| Human Epidermal growth factor Receptor 3 (HER3) | Anti-ErbB3/HER3 antibody [SP71] ab93739 | abcam | 1:100 | Membrane, Cytoplasm |
| Mesenchymal-Epithelial Transition factor (MET) | anti-Total c-MET (SP44) | Ventana | RTU | Membrane, Cytoplasm |
| MUCin-16 (MUC16) | CA-125 (OC125), 05267269001 | Cell Marque Corporation | RTU | Membrane |
| TROPhoblast cell-surface antigen 2 (TROP2) | Anti-TROP2 antibody [EPR20043] ab214488 | abcam | 1:1000 | Membrane |

Supplementary Table 4. Dataset configuration at the independent tumor cell level used to develop the universal immunohistochemistry (UIHC) model.

| **Dataset** | **Class** | **Train** | **Tune** | **Internal test** | **Total** |
| --- | --- | --- | --- | --- | --- |
| PD-L1 22C3 lung | TC+ | 107,438 | 28,763 | 13,792 | 149,993 |
|  | TC- | 568,814 | 59,271 | 37,716 | 665,801 |
| PD-L1 22C3 urothelial carcinoma | TC+ | 115,965 | 19,453 | 17,654 | 153,072 |
|  | TC- | 270,251 | 87,915 | 44,106 | 402,272 |
| PD-L1 22C3 breast cancer | TC+ | 95,556 | 19,331 | 9,789 | 124,676 |
|  | TC- | 269,289 | 71,733 | 38,382 | 379,404 |
| HER2 4B5 Breast cancer | TC+ | 255,661 | 70,882 | 40,805 | 367,348 |
|  | TC- | 306,679 | 97,889 | 41,216 | 445,784 |

PD-L1, Programmed Death-Ligand 1; HER2, Human Epidermal growth factor Receptor 2; TC+, positively stained tumor cell; TC-, negatively stained tumor cell

Supplementary Table 5. Dataset configuration at the independent tumor cell level used to test the universal immunohistochemistry (UIHC) model.

|  | Positively stained tumor cell (TC+) | Negatively stained tumor cell (TC−) |
| --- | --- | --- |
| PD-L1 22C3 lung | 13,792 | 37,716 |
| PD-L1 22C3 urothelial carcinoma | 17,654 | 44,106 |
| PD-L1 22C3 breast cancer | 9,789 | 38,382 |
| HER2 4B5 Breast cancer | 40,805 | 41,216 |
| PD-L1 22C3 pan-cancer (biliary tract, colorectum, liver, stomach, prostate, and pancreas) | 7,259 | 34,491 |
| PD-L1 SP142 lung | 3,437 | 20,916 |
| Claudin 18.2 | 1,173 | 5,614 |
| MET | 1,110 | 5,614 |
| TROP2 | 2,423 | 2,917 |
| MUC16 | 3,464 | 4,043 |
| DLL3 | 974 | 5,664 |
| FGFR2 | 1,181 | 4,193 |
| HER3 | 650 | 3,916 |

PD-L1, Programmed Death-Ligand 1; HER2, Human Epidermal growth factor Receptor 2; MET, Mesenchymal-epithelial transition factor; TROP2, Trophoblast cell-surface antigen 2; DLL3, Delta-like 3; FGFR2, Fibroblast growth factor receptor 2; HER3, Human epidermal growth factor receptor 3.
